# HD-DRUM rhythmic training in Huntington’s disease: feasibility, motor, and brain microstructural pilot signals

**DOI:** 10.64898/2026.01.26.26344842

**Authors:** Pedro Luque Laguna, Vasileios Ioakeimidis, Cheney J. G. Drew, Philip Pallmann, Monica Busse-Morris, Guy B. Watson, Robin Schubert, Anne E. Rosser, Claudia Metzler-Baddeley

**Author notes:** **Corresponding author’s email address**.

## Abstract

**Background:** Rhythmic movement training may benefit cognitive and motor functions in Huntington’s disease (HD). HD-DRUM is a tablet-based application for the training of paced movements. Feasibility of 8 weeks at-home HD-DRUM compared with usual-activity control was assessed in a two-arm, pilot randomised controlled trial (RCT) in people with HD. HD-DRUM-related changes in motor and cognitive functions, mood, and brain microstructure were explored.

**Objectives:** Primary outcomes were feasibility (recruitment, retention, adherence, acceptability) rates for an effectiveness RCT. Secondary outcomes were estimates of HD-DRUM-related effect sizes on motor functions, cognition, mood, and brain microstructure to explore training mechanisms.

**Methods:** Fifty-five individuals with HD were randomised into either HD-DRUM (n = 32, 1 excluded) or usual-activity control (n = 23) groups. Participants underwent Quantitative-Motor, executive function and mood assessments and diffusion-weighted MRI at baseline and 8 weeks follow-up. Recruitment, retention, and adherence (frequency and duration of HD-DRUM engagement) rates were recorded, and HD-DRUM acceptability was assessed with a self-report questionnaire. Group differences of changes in behavioural and brain microstructural measures were reported with 95% confidence intervals.

**Results:** Acceptability of HD-DRUM, retention, and adherence rates averages were high (>80%) and target recruitment was reached. Performance in paced tapping and brain microstructure in motor networks signalled benefits from HD-DRUM and were associated with each other.

**Conclusions:** Acceptability feasibility of HD-DRUM and a future effectiveness RCT in people with HD was established. HD-DRUM-induced brain plasticity may underpin motor performance benefits. HD-DRUM may provide an accessible home-based intervention for maintenance of movements and brain microstructure in HD.

**Trial registration number:** ISRCTN11906973 (Protocol version 1.7, 01/11/2023)

## Introduction

Huntington’s disease (HD) is a neurodegenerative disease caused by a genetic mutation in the *huntingtin* gene and is characterised by the progressive decline in cognition, motor function, and mood. Striatal atrophy [1] and white matter microstructural alterations [2] can be detected many years before disease onset, leading to clinical symptoms [3–5] with debilitating effects on a person’s quality of life [6].

There are no licensed disease-modifying treatments for HD. Although a range of novel therapeutic approaches are under investigation, including gene and cell therapies, with the potential to slow disease progression [7–8], many are complex to deliver, costly, and unlikely to be accessible to all patients. Available symptomatic pharmacological treatments are typically limited in effectiveness and often associated with significant side-effects.

Preclinical studies have shown that environmental enrichment in HD mouse models can slow disease progression, rescue early protein deficits [9–11], and improve the morphology and function of striatal grafts [12,13]. These findings suggest that behavioural or lifestyle interventions have the potential to help with the management of disease progression in HD.

Music-based Rhythmic Auditory Stimulation (RAS) uses external rhythmic cues to facilitate movement [14] and has been shown to improve gait and mobility in people with Parkinson’s disease (PD) [15–17]. Although PD and HD are clinically distinct, both involve basal ganglia (BG) neurodegeneration and impairments in response timing and rhythm generation [18].

The BG, together with auditory and motor cortical regions, form cortico-subcortical networks that support rhythm processing [19, 20]. Key regions implicated in beat-perception include the putamen for generating temporal predictions, the Rolandic operculum for sensorimotor integration, and precentral sulcus and supplementary motor area (SMA) involved in motor planning and control [21]. RAS may therefore compensate for impaired BG timing signals in PD and HD through external rhythmic cueing [14]. However, the neural mechanisms and clinical effects of RAS in HD remain largely unexplored.

Previously, we observed beneficial signals in motor and cognitive function, and brain microstructure following 8-weeks of Bongo drumming training in people with HD [22,23]. We subsequently designed a tablet-based RAS drumming application, HD-DRUM [24], which involves the learning of rhythmic patterns through tapping, while measuring performance and adherence. As a first step in evaluating HD-DRUM as a therapeutic intervention, we conducted a two-arm pilot RCT. The primary objective was to assess the feasibility of a larger effectiveness trial by examining recruitment, retention, adherence and acceptability of 8-weeks home-based HD-DRUM training. Secondary objectives were to estimate effect sizes associated with changes in cognitive and motor performance and MRI-derived microstructural measures in brain regions critical for beat-based rhythmic processing.

## Methods

The study received ethical approval from the Wales Research Ethics Committee 2 (REC reference: 22/WA/0147). It was sponsored by Cardiff University (SPON1895-22) and prospectively registered with ISRCTN 11906973 (https://doi.org/10.1186/ISRCTN11906973). The design was a multi-site two-arm unblinded pilot RCT with parallel HD-DRUM intervention and usual-activity control groups. The protocol including details of the study design, randomisation procedures, outcome measures, and data collection, management, and monitoring plans has been published [25].

### Participants and study setting

Participants were identified, approached, and recruited at five specialist Enroll-HD [26] clinics in the UK (Cardiff, Liverpool, Exeter, Bristol, Birmingham). Participants provided written informed consent prior to taking part in the study.

### Inclusion criteria

Individuals over the age of 18 years with HD as confirmed by genetic testing for the presence of a CAG repeat length ≥ 36 in the huntingtin gene and/or clinical diagnosis, and a Unified Huntington’s Disease Rating Scale (UHDRS) [27] Total Functional Capacity (TFC) score between 9 and 13 (normal function to mild functional impairment).

### Exclusion criteria

A history of any other neurological condition. MRI contra-indications (e.g., pacemakers, stents) precluded MRI assessment but not participation in the trial.

### Study procedure

Participants completed baseline behavioural and MRI assessments at the Cardiff University Brain Research Imaging Centre (CUBRIC) before being randomised to either the HD-DRUM intervention or usual-activity control group. Randomisation was stratified by UHDRS-TFC score to balance disease burden between groups; full details of the randomisation procedure are reported elsewhere [25]. HD-DRUM participants received a Samsung Galaxy 8 tablet with the app and instructions for home use, with researcher support to promote adherence. Control participants continued their usual activities without additional contact. After eight weeks, all participants were invited for follow-up assessments, and HD-DRUM participants completed an acceptability questionnaire. Control participants were offered access to HD-DRUM after completing the study.

### HD-DRUM Intervention

Details of the HD-DRUM content and delivery have been published elsewhere [24]. HD-DRUM comprises 22 audio-guided training sessions introducing rhythmic patterns (e.g. paradiddles, hip-hop, funk, reggaeton) with and without metronome or musical background. Participants were asked to tap along to the rhythms using the left hand on a blue triangle and the right hand on a red circle displayed on the tablet, receiving visual and auditory feedback. Tempo and complexity increased across sessions, with practice progressing from single-hand to bimanual tapping. Participants were instructed to advance at their own pace, repeating sessions as desired. They had to achieve 60-70% response accuracy to progress to the next level. The app recorded response speed, accuracy, and training frequency and duration as adherence measures. Participants received an instruction manual recommending 10-15 minutes of practice, 5 times per week for 8 weeks (40 sessions in total).

### Primary feasibility outcome measures

*Recruitment* was assessed as the proportion of eligible, approached participants who consented and the extent to which the target sample size of 50 was achieved. *Retention* was defined as the proportion of recruited participants who completed the study with reasons for withdrawal and loss to follow-up recorded where possible.

*Adherence* was monitored within HD-DRUM by recording training frequency and duration. Adherence by frequency was calculated as the percentage of training days completed (≥5 minutes per day) relative to the prescribed 40 days (5 days/week for 8 weeks). Adherence by duration was calculated as the percentage of total training minutes completed relative to the expected dose (451 minutes; 40 sessions x 11.3 minutes average session duration).

*Acceptability* was assessed using a self-report questionnaire covering enjoyment, engagement, ease of use, appropriateness of training schedule, perceived effects, likes and dislikes and suggestions for improvements (see Supplementary Material). Overall acceptability was defined as the proportion of participants who agreed with the statement “Overall, I enjoyed using the app and learning to drum along to different rhythms.”

Feasibility success criteria were predefined using a traffic lights system: green (all metrics ≥ 70%) signalling trial success, amber (no metric <40% but at least one <70%) indicating the need for further process evaluation; and red (at least one metric < 40%) indicating that a future RCT would not be feasible.

### Secondary outcome measures

Cognitive and motor assessments were conducted at baseline and follow-up in a quiet testing room at CUBRIC and lasted approximately two hours, including breaks.

#### Cognitive function

Executive function [28] was assessed with paper-and-pencil and computerised tests (from the Psychology Experiment Building Language (PEBL) battery [29] and implemented in MATLAB [30]) including Stroop [31], verbal trails [32], verbal fluency [33], Symbol Digit Modality Test (SDMT) [34], PEBL-Eriksen flanker [35], and MATLAB-based single- and dual-task visual and auditory n-back paradigms [36,37]. Thirty-one cognitive variables were selected for analyses, comprising total correct responses (Stroop, verbal fluency, SDMT), response times (flanker, Stroop, verbal trails), and the sensitivity index A’ [38] for n-back performance.

#### Motor function

Motor performance was assessed with the Quantitative Motor (Q-Motor) test battery [39], including speeded and metronome-paced tapping tasks for left and right index fingers and feet, and dual finger pointing-and-tapping tasks. Metronome tapping was performed at slow and fast tempi, with and without auditory cue. For each condition, mean and standard deviation (SD) were computed for tapping speed (inter-onset interval, IOI; s), force production (area under the force–time curve, AUC) and pacing accuracy (absolute deviation from target tempo). Additional kinematic measures were extracted for speeded and pointing tasks [25, 39]. This yielded 96 measures for metronome tapping, 72 for speeded tapping and 22 measures for the pointing-and-tapping task per session.

#### Mood

Mood was assessed with the Patient Health Questionnaire (PHQ-8) [40].

#### MRI brain microstructure

Multi-shell high angular resolution diffusion imaging (msHARDI) [41] data were acquired on a 3T Siemens Connectom system with strong (300mT/m) gradients [42] using single-shot spin-echo EPI (TR/TE = 3000/59 ms; 2 mm isotropic; GRAPPA=2; AP phase-encoding).

Diffusion weighting comprised b-values of 200, 500, 1200, 2400, 4000 and 6000 s/mm² (20, 20, 30, 61, 61 and 61 directions, respectively), with 15 b=0 s/mm² images (13 AP interleaved plus 2 PA) (∼18 min acquisition time). Additional myelin-sensitive sequences (quantitative magnetisation transfer [43], multicomponent relaxation with steady-state imaging [44]) were acquired but are not reported here.

Diffusion data were pre-processed using MRtrix3 [47] and FSL 6.03, including Marchenko–Pastur PCA denoising [45], Gibbs-ringing correction [46], brain masking, susceptibility-induced distortion correction with TOPUP (AP/PA b=0 images) [48], and motion and eddy-current distortions correction with slice-wise outlier replacement using EDDY [49,50]. Gradient non-linearity [51] and MRI signal-drift corrections were applied using in-house MATLAB scripts.

Four biophysical diffusion models were fitted to derive parametric maps of grey and white matter microstructure. Diffusion tensor imaging (DTI) [52, 53] was fitted to the b = 1200 s/mm² shell to estimate fractional anisotropy (FA), mean diffusivity (MD), and radial diffusivity (RD). Neurite orientation dispersion and density imaging (NODDI) [54] was fitted using b = 200-2400 s/mm² to derive neurite density index (NDI), orientation dispersion index (ODI) and isotropic signal fraction (ISOSF). The Soma and Neurite Density Imaging (SANDI) model [56] was applied to all shells to derive neurite and soma signal fractions (fneurite, fsoma), apparent soma radius (Rsoma), and extracellular diffusivity (De) and the CHARMED model [55] was fit to estimate restricted signal fraction (RF). Parametric maps underwent visual quality control.

Final outcomes were computed by averaging each metric within anatomically defined region of interests (ROIs). Grey matter ROIs were derived using FreeSurfer’s longitudinal stream [57], yielding bilateral masks for the caudate, putamen, globus pallidus, thalamus, primary motor cortex, and SMA areas (paracentral and superior frontal labels from the Desikan-Killiany parcellation). Native-space diffusion maps were projected to each participant’s longitudinal T1 template and averaged within ROIs. White-matter tracts were reconstructed using TractSeg [58, 59] applied to fibre orientation distribution peaks generated in MRtrix3 [47]. Tracts of interest included seven corpus callosum (CC1-7) segments [60], bilateral corticospinal tracts (CST), and striato-prefrontal (ST_PREF) and striato-premotor (ST_PREM) tracts, used as proxies for caudate-prefrontal and putamen-SMA connections. Tract-average metrics were computed with streamline density weighting. In total, 168 grey-matter and 156 white-matter microstructural outcome measures were obtained per participant and session.

### Disease stage classification

Participants’ HD-ISS staging [61] was determined using the online HD-ISS calculator [62]. Genetic and clinical data were obtained from Enroll-HD [26] and normalised striatal volumes were derived from the study-acquired T1-weighted MRI scans.

### Statistical methods

A pragmatic sample size of 50 participants was targeted, allowing feasibility rates to be estimated with a 95% binomial confidence interval (CI) within ± 15 percentage points, irrespective of the observed proportion [63, 64].

#### Feasibility rates

Recruitment and retention data were collected and managed using Research Electronic Data Capture (REDCap) [65, 66]. Adherence was recorded in the HD-DRUM app. Acceptability rating and free-text responses were obtained via self-report questionnaire. Feasibility outcomes were evaluated against the predefined success criteria described above.

#### Principal component analyses

Principal component analysis (PCA) was used to reduce the dimensionality of cognitive, motor, and MRI microstructural outcomes. Percentage change (%Δ) from baseline to follow-up was calculated for each outcome [%Δ = 100 x (Follow-up − Baseline)/Baseline] and separate PCAs were applied to each outcome domain. A single PCA was conducted on %Δ of the 31 cognitive variables (Cog-PC). Four PCAs were conducted on motor outcomes: metronome tapping performance (48 mean variables) (MTM-PC), metronome tapping variability (48 SD variables) (MTV-PC), speeded tapping and pointing performance (47 mean variables) (STM-PC), and variability (47 SD variables) (STV-PC). Two further PCAs were applied to %Δs of grey-matter (168 variables) and white-matter (156 variables) microstructural outcomes (GMM-PC, WMM-PC).

To limit over-extraction in this relatively small sample, component retention followed recommended guidelines [67, 68], applying the Kaiser criterion (eigenvalue > 1) followed by inspection of Cattell scree plots [69]. Retained components were orthogonally rotated using Varimax with Kaiser normalisation [70], and component scores of percentage-changes were calculated for each participant.

#### Estimation of effect sizes of HD-DRUM on secondary outcomes

Effect sizes of the HD-DRUM intervention on secondary outcomes were estimated using par-qcipants’ PCA scores and general linear models (GLMs) for each component, with group (HD-DRUM versus control) as the explanatory variable and age and sex as covariates. Extreme outliers were excluded from each GLM using the Tukey criterion [71], defined as val-ues exceeding three times the interquartile range. Standardised group effect sizes and 95% CIs were displayed using forest plots [72]. For components where the 95% CI did not include zero, violin plots [73] visualised and aided interpretation of intervention effects. Interpreta-qon was guided by the rotated PCA loading pa∼erns, with variables showing absolute loadings ≥ 0.50 considered salient.

#### Correlations between motor and brain microstructure changes

To explore whether HD-DRUM-related changes in motor performance were associated with changes in brain microstructure, we conducted correlation analyses between participants’ scores on motor and brain microstructure PCA components for which the 95% CI of the group effect excluded zero. Pearson and Spearman correlation coefficients were calculated for each motor-microstructural component pair, separately for each group. Results were considered exploratory and reported with uncorrected p-values.

PCAs were conducted in IBM SPSS Statistics (v27) [74]. Descriptive statistics, GLMs, and figures were generated in Python [75] using *statsmodels* [76] and *matplotlib* [77].

#### Mood

Means and SDs of PHQ-8 baseline and follow-up scores were calculated and reported descriptively for each group.

## Results

### Recruitment

Fifty-five participants were recruited, exceeding the target sample size of 50, and representing 35% of approached individuals (Figure 1, Table 1). Reasons for non-enrolment are summarised in Figure 1. One participant initially allocated to HD-DRUM, was excluded due to a TFC score <9, leaving 54 participants for analyses (HD-DRUM: n = 31; control: n = 23). Their demographic and clinical characteristics are summarised in Table 2. Groups were balanced for sex and TFC, but control participants were on average 6.7 years younger. HD-ISS staging was completed in 39 participants (72%); staging was not possible in the remainder due to missing clinical and/or imaging data, CAG repeat length <40, or non-sequential staging (e.g., clinical measures indicating functional before motor impairment). Stage distributions are shown in Table 2.

**Figure 1.**
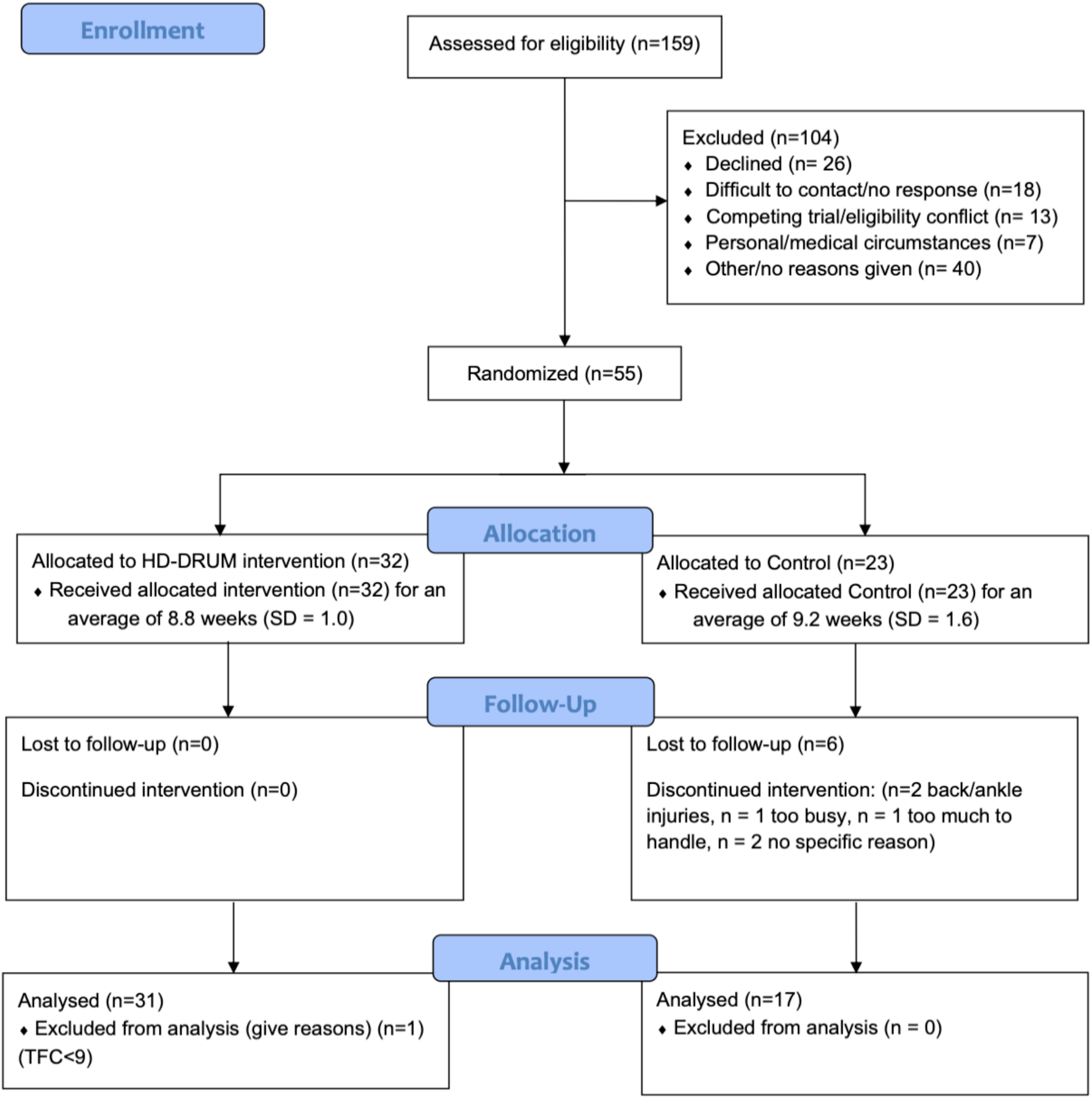
Consolidated standards of reporting trials (Consort) flow chart providing a schematic overview of enrolment, allocation, follow-up, and analysis stages of two-arm randomised controlled feasibility trial of HD-DRUM.

**TABLE 1.** Summary of the feasibility outcomes.

|  | Total HD | HD-DRUM | Control |
| --- | --- | --- | --- |
| Recruited out of approached, n/N (%) | 55/156 (34.6%) | 32 | 23 |
| Recruitment of eligible target sample size, n/N (%) | 54/50 (108%) | 31 | 23 |
| Retained at follow-up, n/N (%) | 48/54 (88.9%) | 31/31 (100.0%) | 17/23 (73.9%) |
| Adherence frequency rate, mean% (range) | - | 81% (5%–145%) | - |
| Adherence frequency rate, <40% / 40%-70% / >70% | - | 4 / 4 / 23 | - |
| Adherence duration rate, mean% (range) | - | 119% (14.8%–348.1%) | - |
| Adherence duration rate, <40% / 40%-70% / >70% | - | 2 / 6 / 23 | - |
| Acceptability (% agreement) | - | 93.1% | - |

Abbreviations: HD = Huntington's disease.

**TABLE 2.** Demographic and clinical background information for eligible participants.

|  | Total HD | HD-DRUM | Control | Control (retained) |
| --- | --- | --- | --- | --- |
| ***Participants characteristics*** |  |  |  |  |
| Group size, n | n = 54 | n = 31 | n = 23 | n = 17 |
| Females, n (%) | 29 (53.7%) | 17 (54.8%) | 12 (52.2%) | 8 (47.1%) |
| Age-years, mean (SD) | 47.8 (12.9) | 50.6 (11.1) | 43.9 (14.1) | 41.4 (14.9) |
| Education-years, mean (SD) | 14.2 (2.7) | 14.8 (2.8) | 13.3 (2.5) | 13.4 (2.6) |
| CAG repeat length, mean (SD) | 42.0 (2.6) | 41.3 (2.2) | 43.3 (2.6) | 43.6 (3.0) |
| TFC, mean (SD) | 11.9 (1.3) | 12.2 (1.2) | 11.5 (1.4) | 11.5 (1.4) |
| SDMT, median (IQR) | 43 (30 – 57) | 47.5 (35.8 – 58) | 34 (26 – 45) | 34 (23.5 – 47.5) |
| TMS, median (IQR) | 9 (3.0– 17.8) | 6.5 (2.25 – 16.5) | 11 (3.75 – 18.5) | 12 (4.5 – 17.8) |
| IS, median (IQR) | 100 (90 – 100) | 100 (95 – 100) | 95 (85 – 100) | 95 (85 – 100) |
| MOCA, mean (SD) | 25.3 (3.6) | 26.2 (3.1) | 24.1 (3.8) | 24.1 (4.2) |
| HD-ISS | n | n | n | N |
| No CAG available | 3 | 0 | 3 | 2 |
| CAG < 40 | 6 | 6 | 0 | 0 |
| Undefined* | 3 | 2 | 1 | 0 |
| Indeterminate 0 vs 1** | 5 | 2 | 3 | 2 |
| HD-ISS = 0 | 4 | 2 | 2 | 2 |
| HD-ISS = 1 | 7 | 5 | 2 | 2 |
| HD-ISS = 2 | 9 | 6 | 3 | 1 |
| HD-ISS = 3 | 17 | 8 | 9 | 8 |

Abbreviations: HD-ISS = Huntington's Disease Integrated Staging System, IS = Independent Scale, MOCA = Montreal Cognitive Assessment, SDMT = Symbol Digit Modality Test, TFC = Total Functional Capacity, TMS = Total Motor Score. \* Undefined due to non-sequential staging,
\*\* Indeterminate stage 0 versus 1 due to MRI biomarker not being available.

### Retention

No participants withdrew from the HD-DRUM group (100% retention) but six participants from the control group (73.9% retention), resulting in an overall retention rate of 88.9% (Figure 1, Table 1). Reasons for withdrawal are summarised in Figure 1.

### Adherence

Mean adherence was 81% for frequency and 100% for duration (Table 1). Most participants (23/31) met the predefined 70% criterion, engaging with HD-DRUM on at least 28 occasions for a total duration of ≥ 315 min; maximum engagement reached 58 sessions (27 h). Six participants showed moderate adherence (40–69%), while two showed minimal adherence (< 40%), including one participant with no meaningful engagement (2 sessions, 1 h total).

### Acceptability

Overall, participants rated the HD-DRUM app as enjoyable (94%), engaging (81%), and easy to use (93.5%), with most valuing the training content, musical elements, and research team support (87%; Supplementary Table 1). Perceived benefits in cognition, motor function, or mood were small-modest (29–48% agreement), although around half reported improvements in hand coordination (55%). A prominent theme was the need for clearer and more personalised performance feedback (77% desired additional in-app feedback). Some participants reported challenges with specific sessions or technical issues (lack of responsiveness, freezing). Thus, HD-DRUM was viewed as highly enjoyable, accessible, and engaging but required improved personalisation, enhanced feedback, and technical refinements to optimise acceptability.

### Effect on cognitive performance

Data from 27 participants (19 HD-DRUM) were included in the PCA due to missing data. Eleven components were identified, explaining 84% of data variance. One extreme outlier was excluded from each of four components. The GLM showed a significant group effect for Cog-PC11, which loaded primarily on response speed in congruent flanker trials (Figure 2). This effect reflected slower baseline performance of control relative to HD-DRUM participants rather than a drumming-related change.

**Figure 2.**
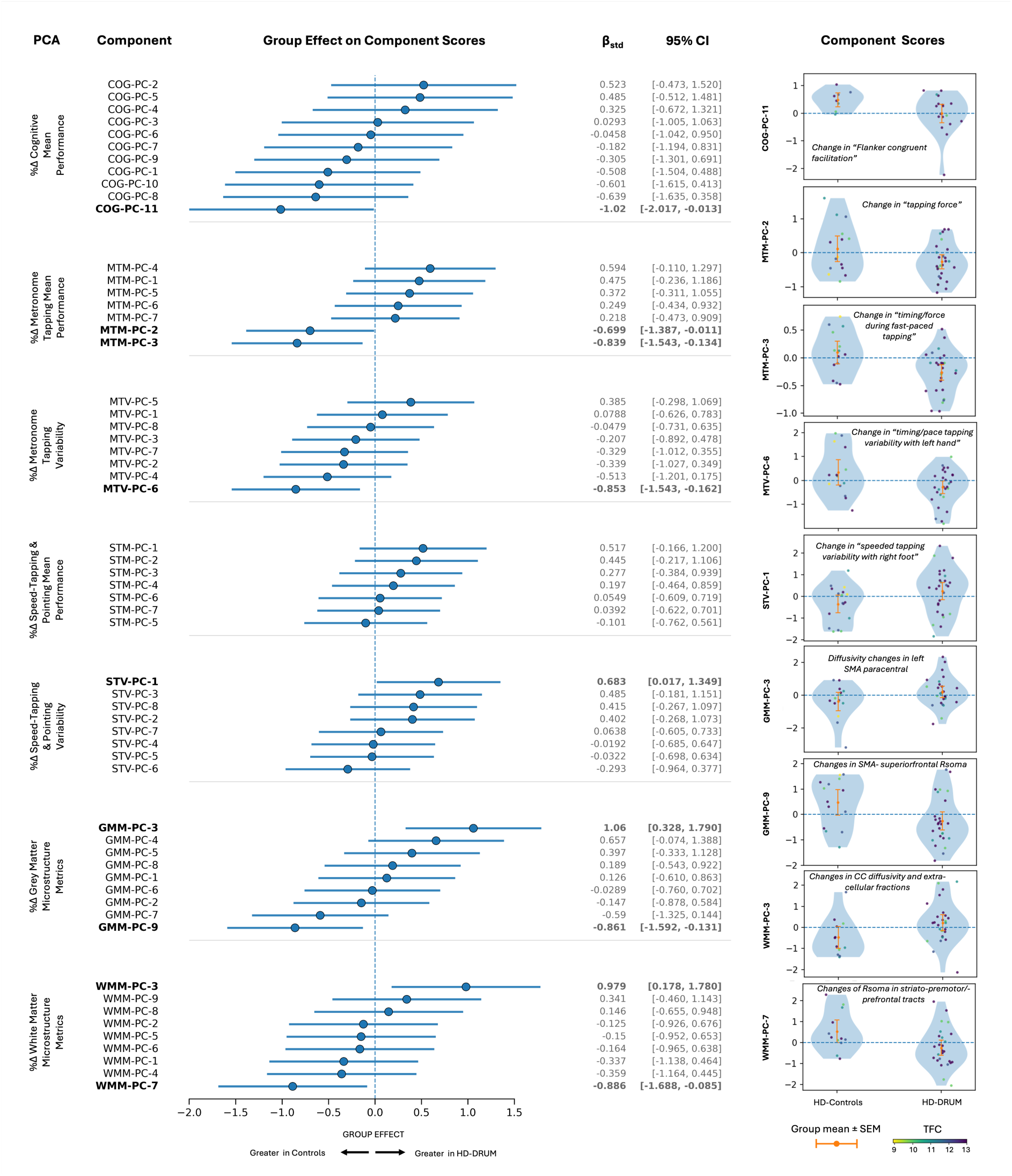
Effects of the HD-DRUM intervention on cognitive performance, motor performance, and brain microstructure. **Left**: Forest plots of standardized between-group effects (β_std_; 95% CI) on principal-component (PC) scores from general linear models, derived from seven separate principal component analyses (PCAs) of percentage longitudinal change (%Δ) in outcome measures of Cognitive Performance (COG), of Metronome Tapping Mean Performance (MTM), of Metronome Tapping Variability (MTV), of Speeded-Tapping and Pointing Mean Performance (STM), of Speeded-Tapping and Pointing Variability (STV), of Grey-Matter Microstructure (GMM), and of White-Matter Microstructure (WMM). All models were adjusted for age and sex, and extreme outliers identified by the Tukey criterion (k = 3 × IQR) were excluded per component. The horizontal Group Effect axis is annotated such that positive β_std_ indicates higher PC scores in HD-DRUM than in Controls. PCs whose 95% CI excludes zero are shown in bold and carried forward to the middle and right columns. **Right**: Violin plots of the score distributions of these selected PCs in HD-Controls and HD-DRUM, with individual participants overlaid (HD-DRUM points colour-coded by Total Functional Capacity, TFC; range 9–13) and group mean ± SEM shown in orange. In-panel descriptors summarise the variables based on the largest absolute rotated loadings on each PC.

### Effect on motor performance

Across the four Q-Motor PCAs, seven or eight components were retained per analysis, explaining approximately 60% of the variance. Thirteen participants (including five controls) were identified as extreme outliers across up to two components each, resulting in fiÅeen GLMs with one exclusion and two GLMs with two exclusions. For four components across three Q-Motor PCAs, the 95% CIs of the group effect size excluded zero (Figure 2). MTM-PC2 and PC3 showed large group effects driven by longitudinal decreases in the HD-DRUM group relative to increases in controls. Salient loadings indicated improvements in tapping force (MTM-PC2) and faster, more controlled timing and force during fast-paced tapping (MTM-PC3) following drumming. MTV-PC6, characterised by leÅ-hand tapping measures, showed reduced variability aÅer HD-DRUM compared with increased variability in controls. Finally, STV-PC1, loading on right-foot speeded tapping, reflected reduced variability in controls but no change in the HD-DRUM group; baseline differences indicated greater variability in controls that resolved at follow-up.

#### Effect on Mood

Mean (SD) PHQ-8 scores were 4.29 (4.79) at baseline and 4.16 (4.38) at follow-up in the HD-DRUM group, and 7.00 (7.25) and 7.64 (7.80) in controls indicating no change in mood in either group.

### Effect on brain microstructure

GMM-PCA included 41 participants (27 HD-DRUM) and WMM-PCA 38 participants (26 HD-DRUM), due to failed tract reconstructions. Both PCAs retained nine components; one extreme outlier was excluded from each of six GLMs. Two GMM-PCA components showed group effects with 95% CIs excluding zero (Figure 2). GMM-PC3, loading on leÅ SMA–paracentral DTI and CHARMED metrics, was associated with decreased MD and RD and increased FA and FR in HD-DRUM, with the opposite pa∼ern in controls. GMM-PC9, loading on bilateral SMA–superior frontal Rsoma, showed small decreases in HD-DRUM versus increases in controls. Two WMM-PCA components also showed group effects with 95% CIs excluding zero. WMM-PC3, loading on central callosal diffusivity, ISOSF, and extracellular-fraction metrics, reflected larger longitudinal decreases in controls and relative stability in HD-DRUM. WMM-PC7, loading on Rsoma in bilateral striato-prefrontal and striato-premotor tracts, increased in controls but not in HD-DRUM. Thus, HD-DRUM-related signals were observed in SMA regions, central/anterior CC, and striato-prefrontal/premotor tracts.

### Correlations between motor and brain microstructure changes

Correlation coefficients were calculated between the Q-Motor (MTM-PC2, MTM-PC3, MTV-PC6, STV- PC1) and brain microstructure components (GMM-PC3, GMM-PC9, WMM-PC3, WMM-PC7) that showed group effect sizes with 95% CIs excluding zero (Figure 3). In the HD-DRUM group (*n* = 24), higher scores on MTV-PC6 (leÅ-hand tapping variability increase, right-hand decrease) were negatively associated with GMM-PC9 (SMA–superior-frontal Rsoma) (Pearson *r* = −0.46, *p* = 0.02; Spearman ρ = −0.45, *p* = 0.03). No significant correla-qons were observed in the control group.

**Figure 3.**
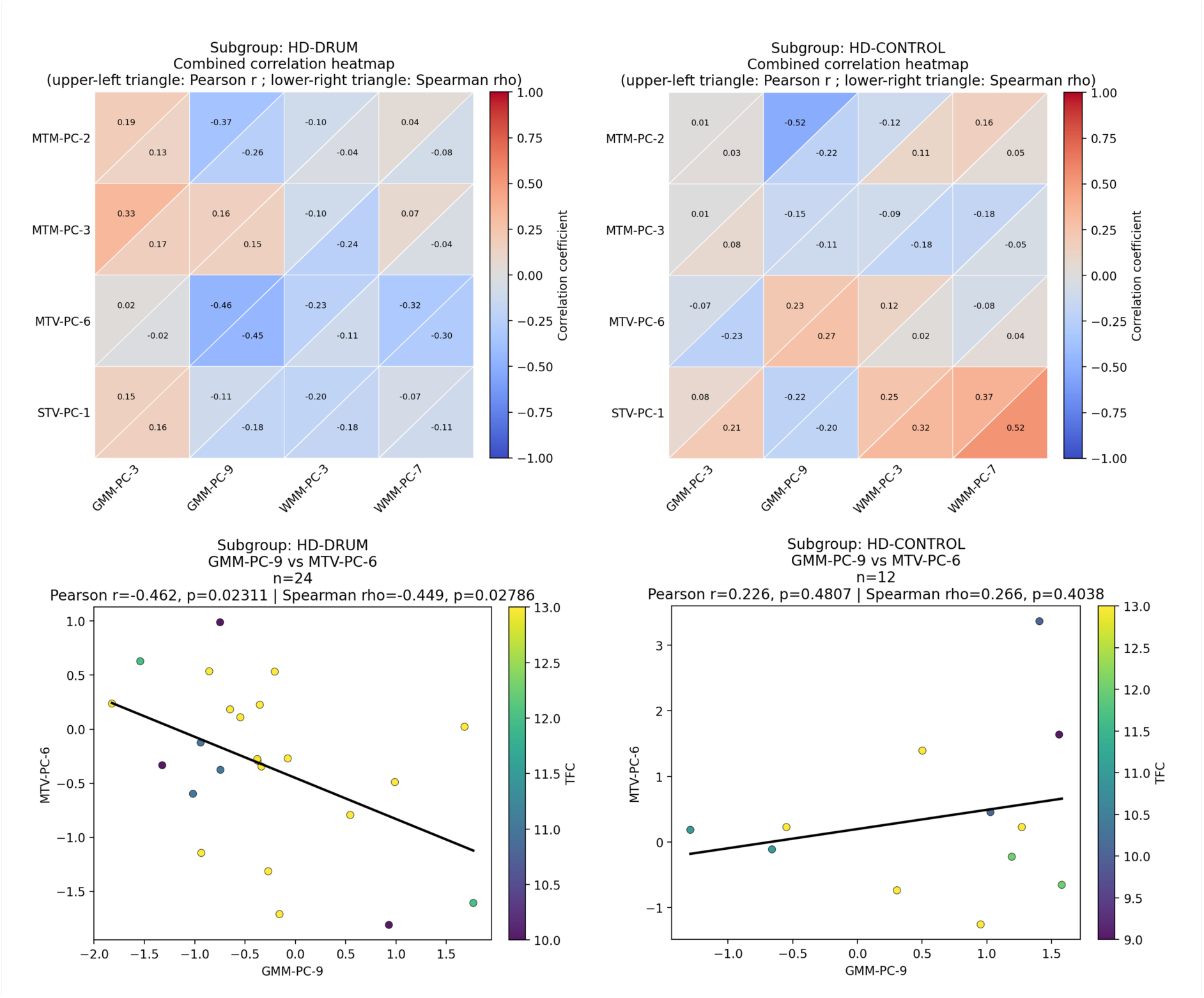
Post-hoc associations between motor and brain microstructure changes. **Top:** Combined correlation heatmaps between the four Q-Motor (rows; MTM-PC-2 and MTM-PC-3 from Metronome Tapping Mean Performance, MTV-PC-6 from Metronome Tapping Variability, and STV-PC-1 from Speeded-Tapping and Pointing Variability) and four brain-microstructure (columns; GMM-PC-3 and GMM-PC-9 from Grey-Matter Microstructure, WMM-PC-3 and WMM-PC-7 from White-Matter Microstructure) principal-component scores selected by the prior GLMs (95% CI excluding zero), shown separately for HD-DRUM (left) and HD-Controls (right). Each cell is split into Pearson r (upper-left triangle) and Spearman ρ (lower-right triangle), both colour-coded on the same [−1, 1] scale. Computations used pairwise-complete observations after per-variable masking of extreme outliers (Tukey k = 3 × IQR). **Bottom:** Scatter plots of the only pair reaching p < 0.05, MTV-PC-6 versus GMM-PC-9, in HD-DRUM (left; n = 24; Pearson r = −0.46, p = 0.023; Spearman ρ = −0.45, p = 0.028) and Controls (right; n = 12; Pearson r = 0.23, p = 0.48; Spearman ρ = 0.27, p = 0.40), with linear fits overlaid. **Abbreviations:** GMM, Grey-Matter Microstructure; HD, Huntington’s disease; IQR, interquartile range; MTM, Metronome Tapping Mean Performance; MTV, Metronome Tapping Variability; PC, principal component; STV, Speeded-Tapping and Pointing Variability; WMM, White-Matter Microstructure.

## Discussion

This pilot RCT evaluated the feasibility and explored preliminary effects of an 8-week, home-based, tablet-delivered rhythmic movement training programme (HD-DRUM) compared with a usual-activity control in individuals with HD. The primary objective was to de-termine whether a larger effectiveness trial would be achievable, assessed through recruit-ment, retention, adherence, and user acceptability, benchmarked against a predefined feasibility threshold of 70%.

All feasibility targets were met. Recruitment exceeded the planned sample size of 50 within the 24-month recruitment window, and overall adherence and retention were high (83%), with no drop-outs in the HD-DRUM group. Participants rated the intervention as highly en-joyable, engaging, and easy to use, with most appreciating the musical content and support provided by the research team. These findings demonstrate that digitally delivered, home-based rhythmic movement training is practical and acceptable for people with HD, and that recruitment, adherence, and retention in a future effectiveness RCT are feasible.

Although participants did not perceive improvements in cognition or mood over the 8-week period, more than half reported improvements in hand coordination. Participant feedback highlighted challenges with training sessions involving irregular rhythms, bimanual coordi-naqon, or reversals, and technical issues such as freezing or reduced responsiveness. Sug-gested improvements focused on be∼er tailoring of training difficulty and clearer, more personalised in-app performance feedback to support progress monitoring. These insights are pivotal for optimising the intervention prior to larger-scale evaluation.

The study also explored HD-DRUM-related signals on motor performance, cognition, mood, and brain microstructure to inform outcome selection, effect size estimation, and potential mechanisms for a future RCT. Exploratory motor analyses suggested that HD-DRUM primarily affected rhythmic paced motor control. Improvements were observed in metro-nome-paced tapping, particularly in components related to timing accuracy, force control, and performance consistency. These effects were most evident during fast-paced tapping and in the leÅ hand, with no corresponding improvements in speeded tapping or dual tap-ping-and-pointing tasks. These findings suggests that 8-weeks of HD-DRUM training specifi-cally enhanced rhythmic sensorimotor synchronisation rather than general motor speed or multitasking.

Improvements were particularly apparent in the leÅ, non-dominant hand for most parqci-pants (29/31 were right-handed). As non-dominant hand motor control is typically less precise and slower than that of the dominant hand [78], it may be especially responsive to re-peqqve rhythmic training. Benefits of rhythmic finger tapping exercises were also found to transfer to foot tapping, likely due to rhythmic entrainment, i.e., the natural human tendency to synchronize finger and foot tapping movements to an external beat [79].

Exploratory neuroimaging findings provided converging evidence for potential neural mech-anisms supporting these behavioural effects. HD-DRUM–related microstructural changes were observed in motor-related cortical and subcortical structures, including the SMA and paracentral cortices and striato-prefrontal and striato-premotor tracts. HD-DRUM-training was associated with relative maintenance or improvements in microstructural indices, whereas controls tended to show changes consistent with disease progression. Importantly, SMA microstructural changes correlated with changes in finger tapping variability, suggesting a link between training-related neural plasticity and improved rhythmic motor control.

These observations are encouraging, as the implicated regions are central to beat percep-qon, predictive timing, and motor coordination [80,81]. Elevated apparent RSoma has previously been observed in the striatum of people with HD compared to healthy controls [82].

Against this background, the relative maintenance of apparent RSoma in the HD-DRUM group raises the possibility that rhythmic training may help slow microstructural decline in motor networks.

As a feasibility study, the trial was not designed or powered to assess clinical efficacy or long-term disease modification. The absence of observed effects on cognition and mood contrasts with earlier uncontrolled pilot work [22,23], likely reflecting the short intervention duration, low baseline mood symptoms, and the importance of controlling for practice ef-fects. In progressive conditions such as HD, benefits may manifest as slowed decline rather than short-term improvement, requiring longer follow-up to detect meaningful effects.

Several considerations emerged for future trials. Only 35% of eligible individuals enrolled, partly due to competition with other clinical trials and time constraints. Drop-out occurred only in the control group, suggesting that an active control condition providing some en-gagement may improve retention. Group imbalances in age, disease stage, and sample size may also have influenced exploratory outcomes and should be addressed in future designs.

In conclusion, HD-DRUM appears feasible, acceptable, and engaging as a home-based rhythmic movement intervention for people with HD. Preliminary findings suggest specific improvements in rhythmic motor control and associated brain microstructure within beat-perception networks. A fully powered RCT with an active control, prespecified primary outcomes, longer follow-up, and an optimised intervention is warranted to determine clinical effectiveness, durability of effects, and impact on disease progression and quality of life.

## Data Availability

All data produced in the present study will be available upon reasonable request to Enroll–HD after completion of the study.

## Data Access

This project was endorsed by the Enroll-HD Scientific Oversight Committee (SOC) (14/11/2022) and used Enroll-HD captured clinical data. At the end of the project, the coded participant level data will be shared and made accessible through the Enroll-HD specific data request process (enroll-hd.org).

## Acknowledgements

We would like to thank the clinical and administerial staff at the participating patient identification centres for their help with identifying suitable patients for the study. We would like to thank Eileen Donovan, Kim Munnery, Jane Davies and Sophie Rowlands from the Cardiff HD clinic; Natalie Rosewell, Anya Soonderpershad, and Professor Elizabeth Coulthard from the Bristol Brain Centre; Dr Timothy Harrower, Claire Tilley, and Jessica Prado Mota from the Royal Devon University Healthcare NHS Foundation Trust in Exeter; Jennifer De Souza and Dr Hugh Rickards from the Birmingham and Solihull Mental Health NHS Foundation Trust; and Dr Sundus Alusi and Jenni Burns from the Walton Centre NHS Foundation Trust in Liverpool. We would like to thank Beate Galoburda, Lucy Layland and Amy Dangerfield for their help with data collection. The scripted pipeline used for the preprocessing of the MRI data was written by Dr Chantal Tax and Dr Greg Parker. In addition, we would like to thank all Public Involvement contributors and the members of the Enroll-HD Scientific Oversight Committee for their input into the study as well as all participants for their generous time commitment to help us assess the feasibility of HD-DRUM.

## Author’s roles

PLL: execution, analysis, writing, editing

VI: execution, editing

MBM: design, editing

CJGD: design, editing

PP: design, editing

GBW: design, editing

RS: design, analysis

AER: design, editing

CMB: design, analysis, writing, editing

## Supplements

**Table S1.**
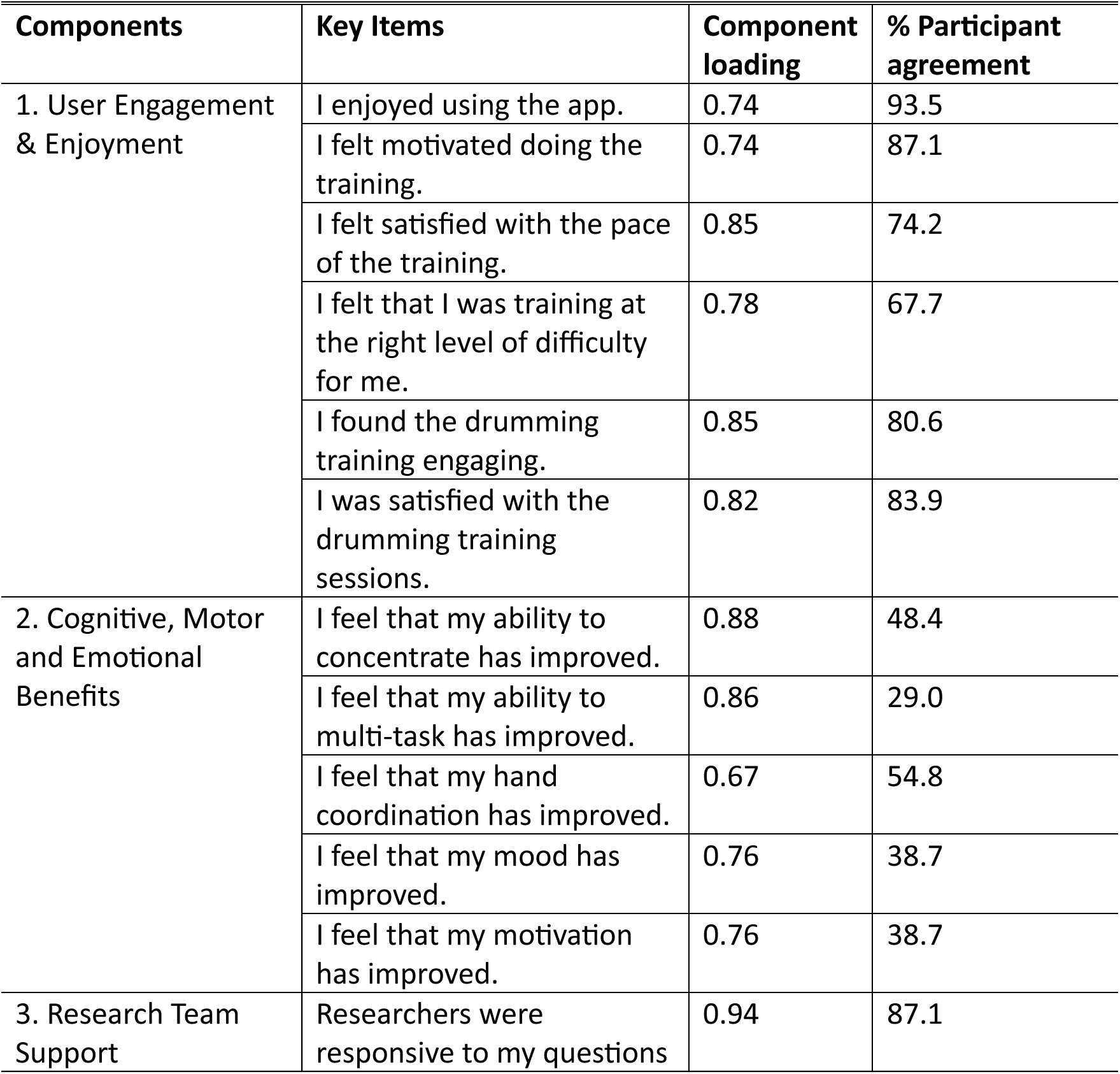
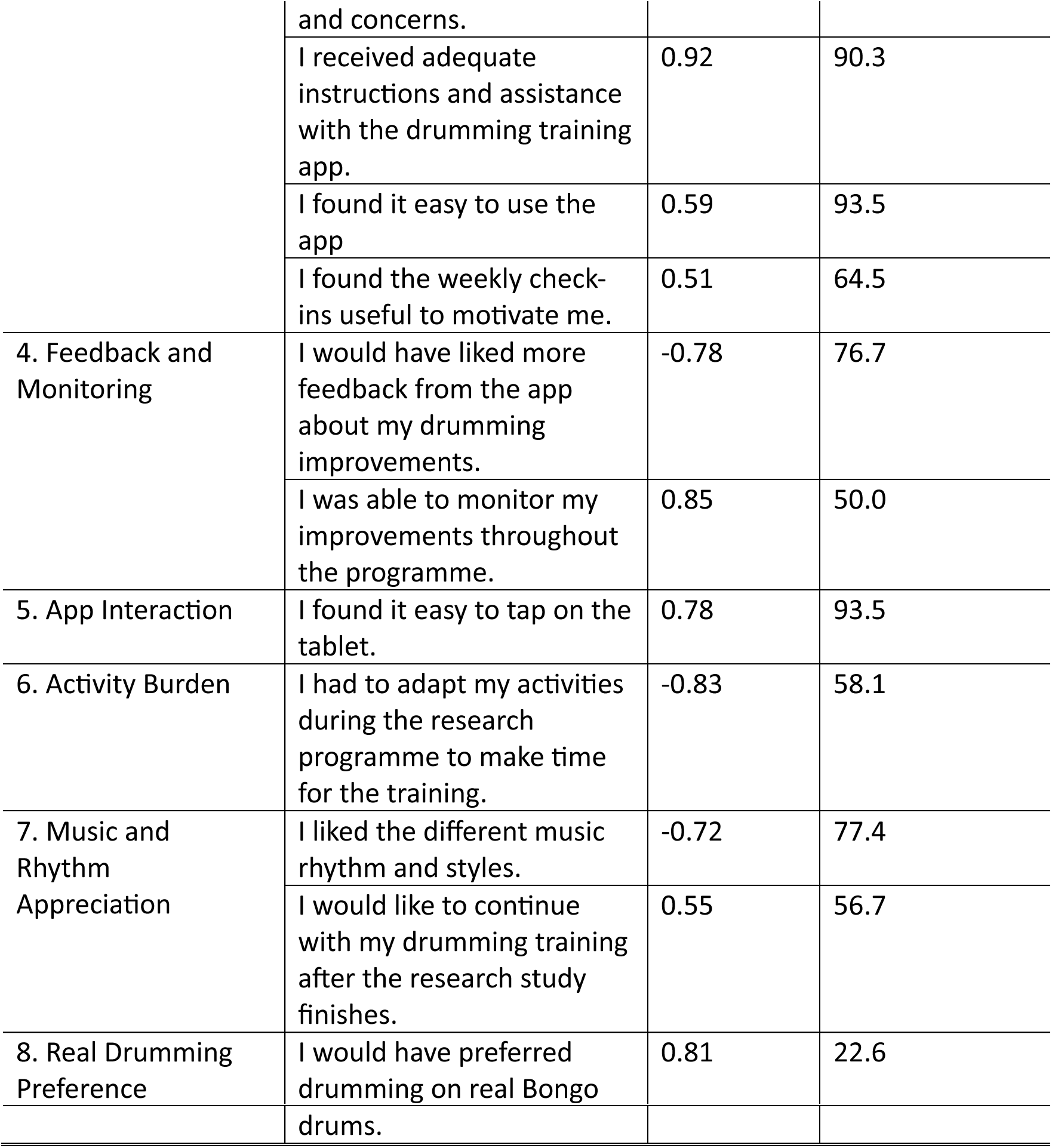
Thematic components and items of acceptability questionnaire.

| Components | Key Items | Component loading | % Participant agreement |
| --- | --- | --- | --- |
| 1. User Engagement & Enjoyment | I enjoyed using the app. | 0.74 | 93.5 |
|  | I felt motivated doing the training. | 0.74 | 87.1 |
|  | I felt satisfied with the pace of the training. | 0.85 | 74.2 |
|  | I felt that I was training at the right level of difficulty for me. | 0.78 | 67.7 |
|  | I found the drumming training engaging. | 0.85 | 80.6 |
|  | I was satisfied with the drumming training sessions. | 0.82 | 83.9 |
| 2. Cognitive, Motor and Emotional Benefits | I feel that my ability to concentrate has improved. | 0.88 | 48.4 |
|  | I feel that my ability to multi-task has improved. | 0.86 | 29.0 |
|  | I feel that my hand coordination has improved. | 0.67 | 54.8 |
|  | I feel that my mood has improved. | 0.76 | 38.7 |
|  | I feel that my motivation has improved. | 0.76 | 38.7 |
| 3. Research Team Support | Researchers were responsive to my questions and concerns. | 0.94 | 87.1 |
|  | I received adequate instructions and assistance with the drumming training app. | 0.92 | 90.3 |
|  | I found it easy to use the app | 0.59 | 93.5 |
|  | I found the weekly check-ins useful to motivate me. | 0.51 | 64.5 |
| 4. Feedback and Monitoring | I would have liked more feedback from the app about my drumming improvements. | -0.78 | 76.7 |
|  | I was able to monitor my improvements throughout the programme. | 0.85 | 50.0 |
| 5. App Interaction | I found it easy to tap on the tablet. | 0.78 | 93.5 |
| 6. Activity Burden | I had to adapt my activities during the research programme to make time for the training. | -0.83 | 58.1 |
| 7. Music and Rhythm Appreciation | I liked the different music rhythm and styles. | -0.72 | 77.4 |
|  | I would like to continue with my drumming training after the research study finishes. | 0.55 | 56.7 |
| 8. Real Drumming Preference | I would have preferred drumming on real Bongo drums. | 0.81 | 22.6 |

**Table S2.**
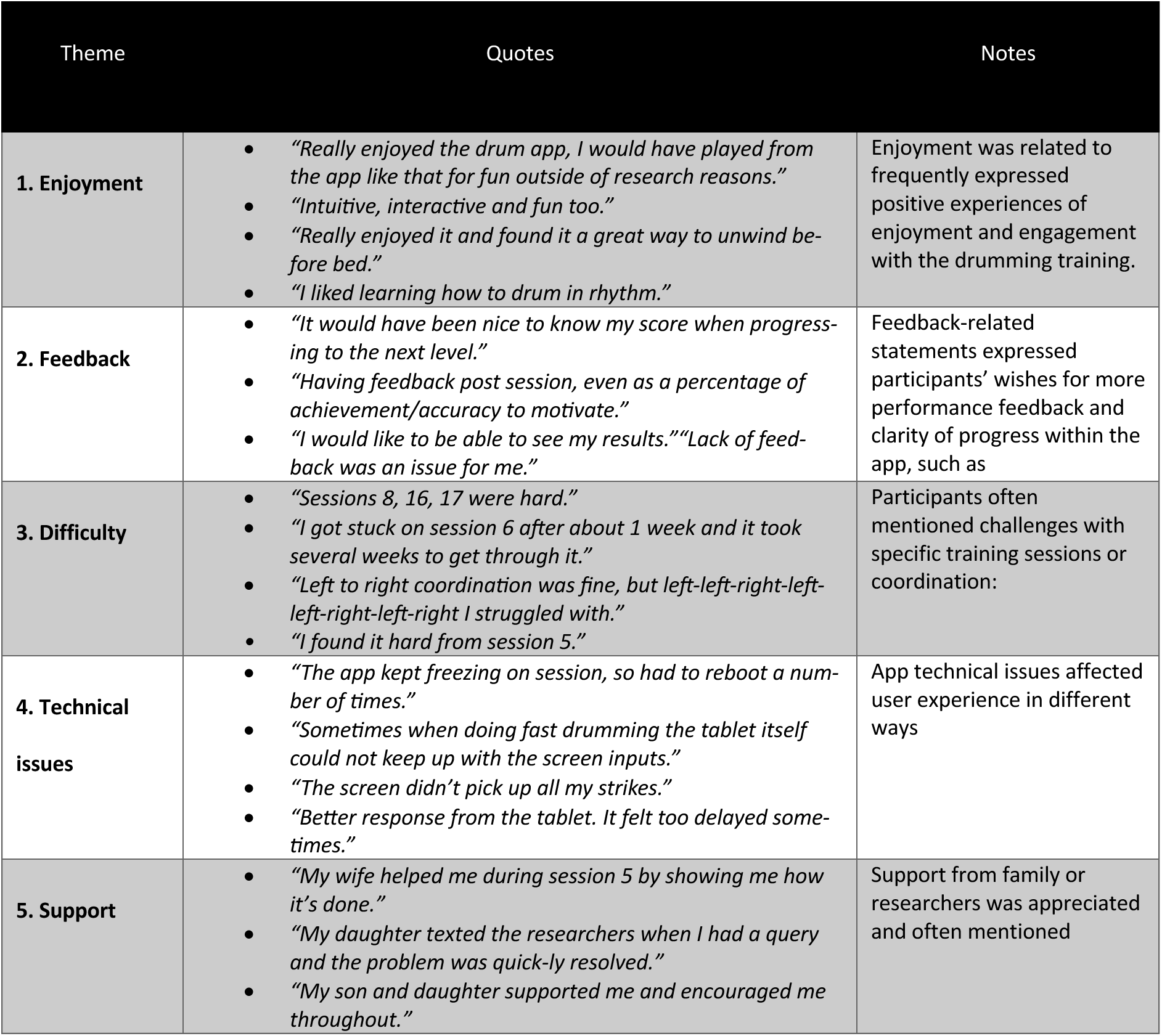
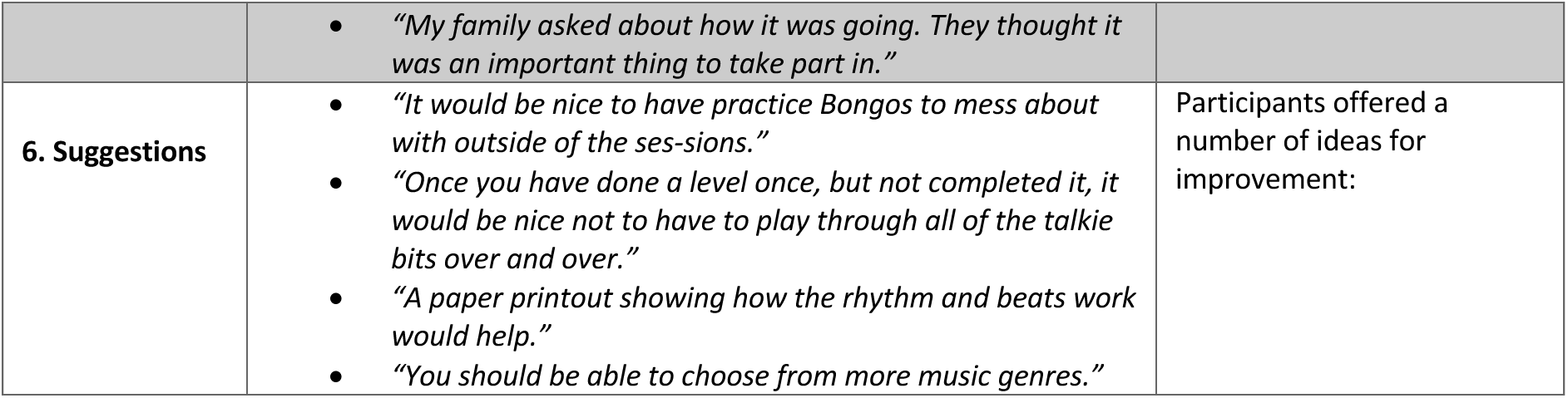
Quotes from thematic analysis of participants’ qualitative feedback.

| Theme | Quotes | Notes |
| --- | --- | --- |
| **1. Enjoyment** | • *"Really enjoyed the drum app, I would have played from the app like that for fun outside of research reasons."*<br>• *"Intuitive, interactive and fun too."*<br>• *"Really enjoyed it and found it a great way to unwind before bed."*<br>• *"I liked learning how to drum in rhythm."* | Enjoyment was related to frequently expressed positive experiences of enjoyment and engagement with the drumming training. |
| **2. Feedback** | • *"It would have been nice to know my score when progressing to the next level."*<br>• *"Having feedback post session, even as a percentage of achievement/accuracy to motivate."*<br>• *"I would like to be able to see my results." "Lack of feedback was an issue for me."* | Feedback-related statements expressed participants' wishes for more performance feedback and clarity of progress within the app, such as |
| **3. Difficulty** | • *"Sessions 8, 16, 17 were hard."*<br>• *"I got stuck on session 6 after about 1 week and it took several weeks to get through it."*<br>• *"Left to right coordination was fine, but left-left-right-left-left-right-left-right I struggled with."*<br>• *"I found it hard from session 5."* | Participants often mentioned challenges with specific training sessions or coordination: |
| **4. Technical issues** | • *"The app kept freezing on session, so had to reboot a number of times."*<br>• *"Sometimes when doing fast drumming the tablet itself could not keep up with the screen inputs."*<br>• *"The screen didn't pick up all my strikes."*<br>• *"Better response from the tablet. It felt too delayed sometimes."* | App technical issues affected user experience in different ways |
| **5. Support** | • *"My wife helped me during session 5 by showing me how it's done."*<br>• *"My daughter texted the researchers when I had a query and the problem was quickly resolved."*<br>• *"My son and daughter supported me and encouraged me throughout."* | Support from family or researchers was appreciated and often mentioned |
|  | • *"My family asked about how it was going. They thought it was an important thing to take part in."* |  |
| **6. Suggestions** | • *"It would be nice to have practice Bongos to mess about with outside of the sessions."*<br>• *"Once you have done a level once, but not completed it, it would be nice not to have to play through all of the talkie bits over and over."*<br>• *"A paper printout showing how the rhythm and beats work would help."*<br>• *"You should be able to choose from more music genres."* | Participants offered a number of ideas for improvement: |

**Figure S1.**
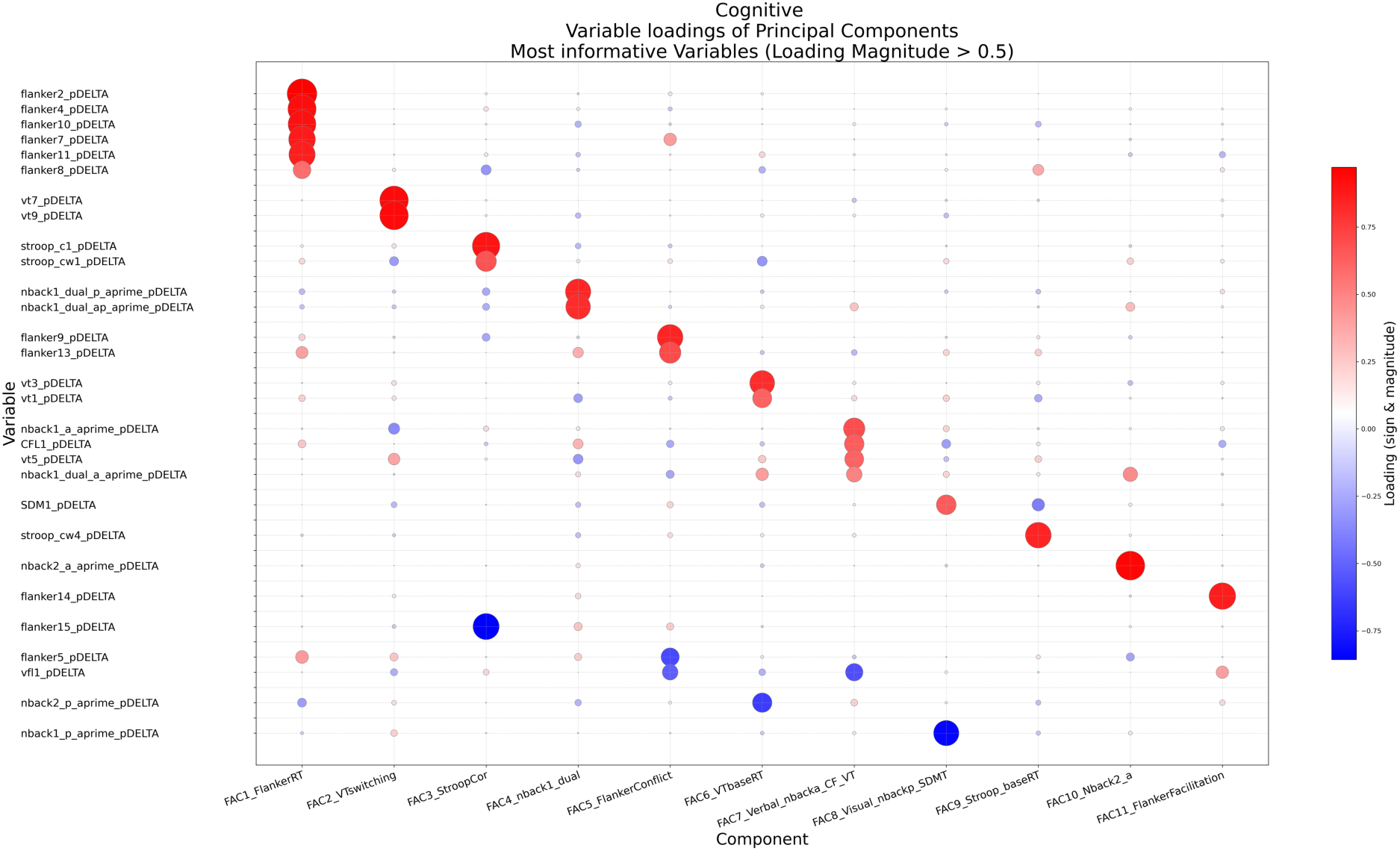
Bubble grid of component loadings for the Cognitive Performance PCA. Rows show total number correct and response time variables included in the Cognitive Performance PCA and columns show retained principal components. Circle area is proportional to the absolute loading magnitude, and colour indicates loading sign and magnitude (red = positive; blue = negative), highlighting variables that contribute most strongly to each component.

**Figure S2.**
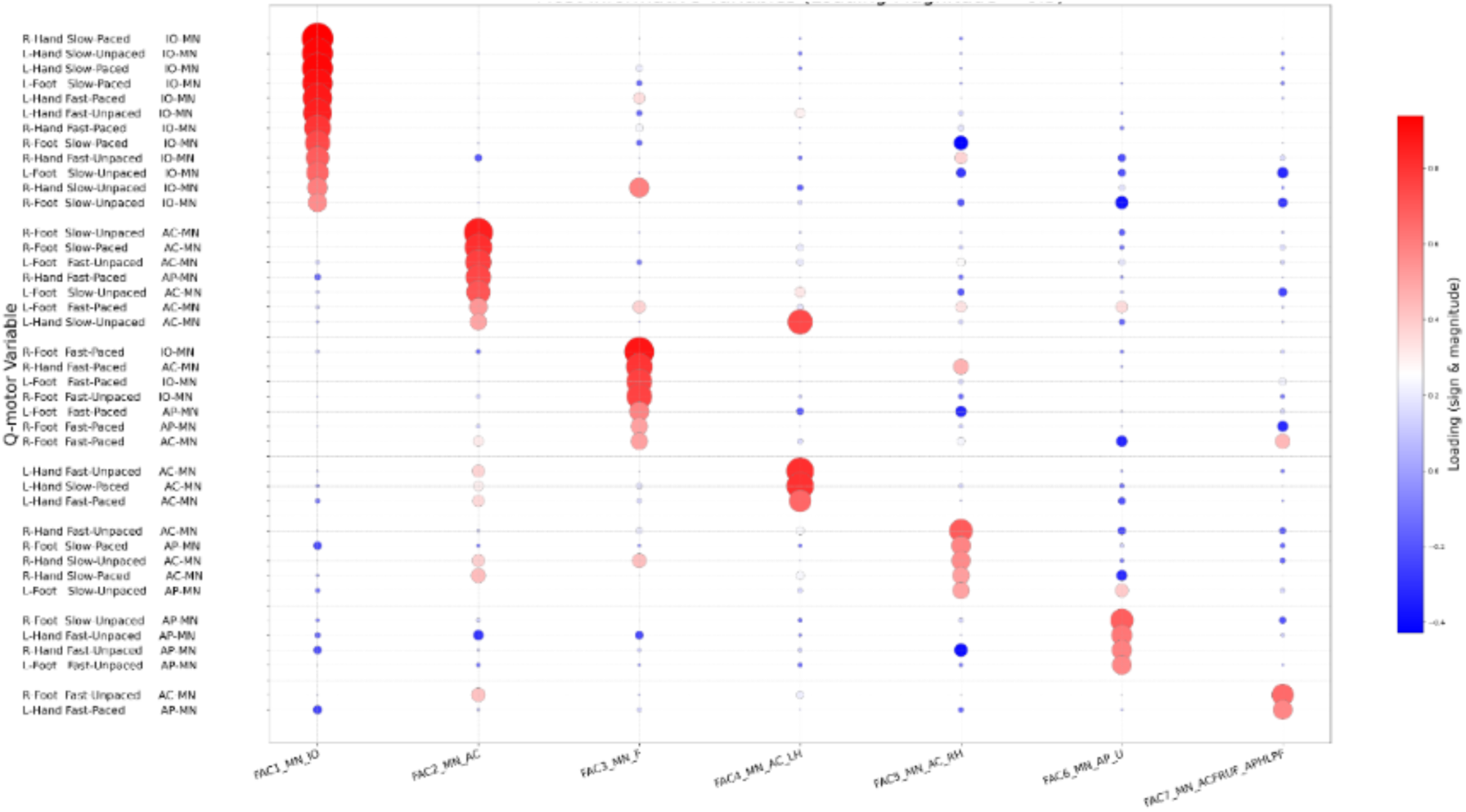
Bubble grid of component loadings for the Metronome Tapping Performance PCA. Rows show “means” variables included in the Metronome Tapping Performance PCA and columns show retained principal components. Circle area is proportional to the absolute loading magnitude, and colour indicates loading sign and magnitude (red = positive; blue = negative), highlighting variables that contribute most strongly to each component.

**Figure S3.**
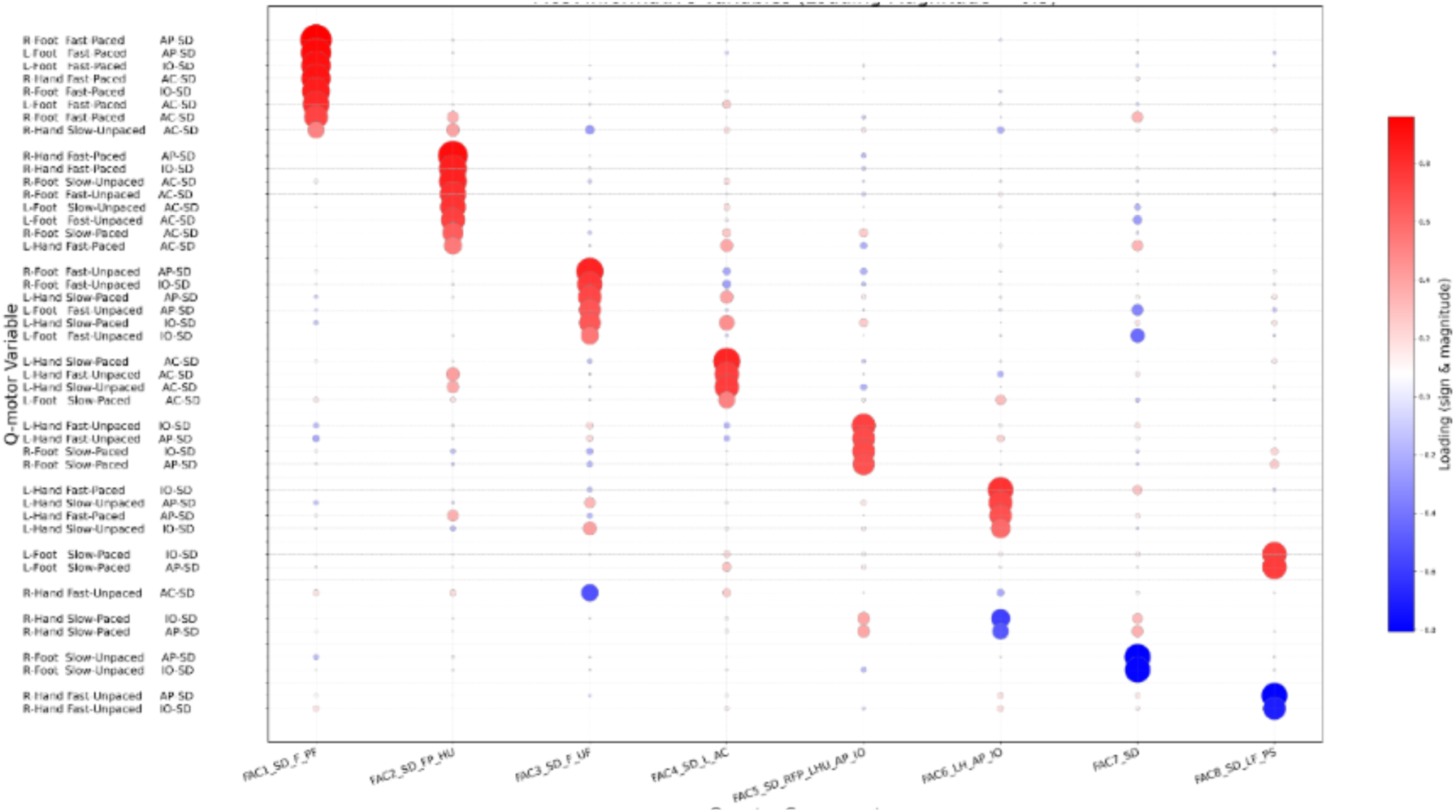
Bubble grid of component loadings for the Metronome Tapping Variability PCA. Rows show “SD” variables included in the Metronome Tapping Variability PCA and columns show retained principal components. Circle area is proportional to the absolute loading magnitude, and colour indicates loading sign and magnitude (red = positive; blue = negative), highlighting variables that contribute most strongly to each component.

**Figure S4.**
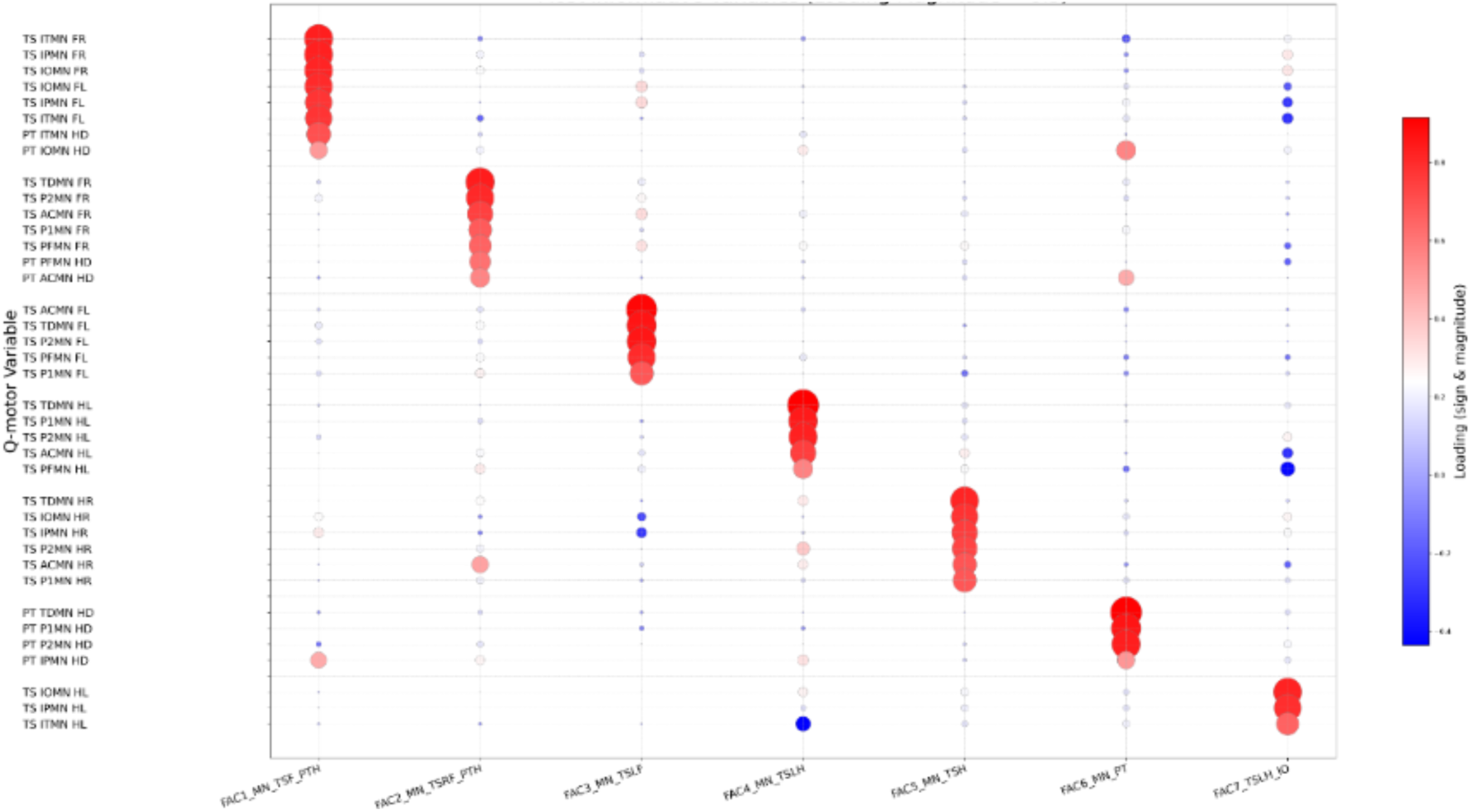
Bubble grid of component loadings for the Speeded Tapping and Pointing Performance PCA. Rows show “mean” variables included in the Speeded Tapping and Pointing Variability PCA and columns show retained principal components. Circle area is proportional to the absolute loading magnitude, and colour indicates loading sign and magnitude (red = positive; blue = negative), highlighting variables that contribute most strongly to each component.

**Figure S5.**
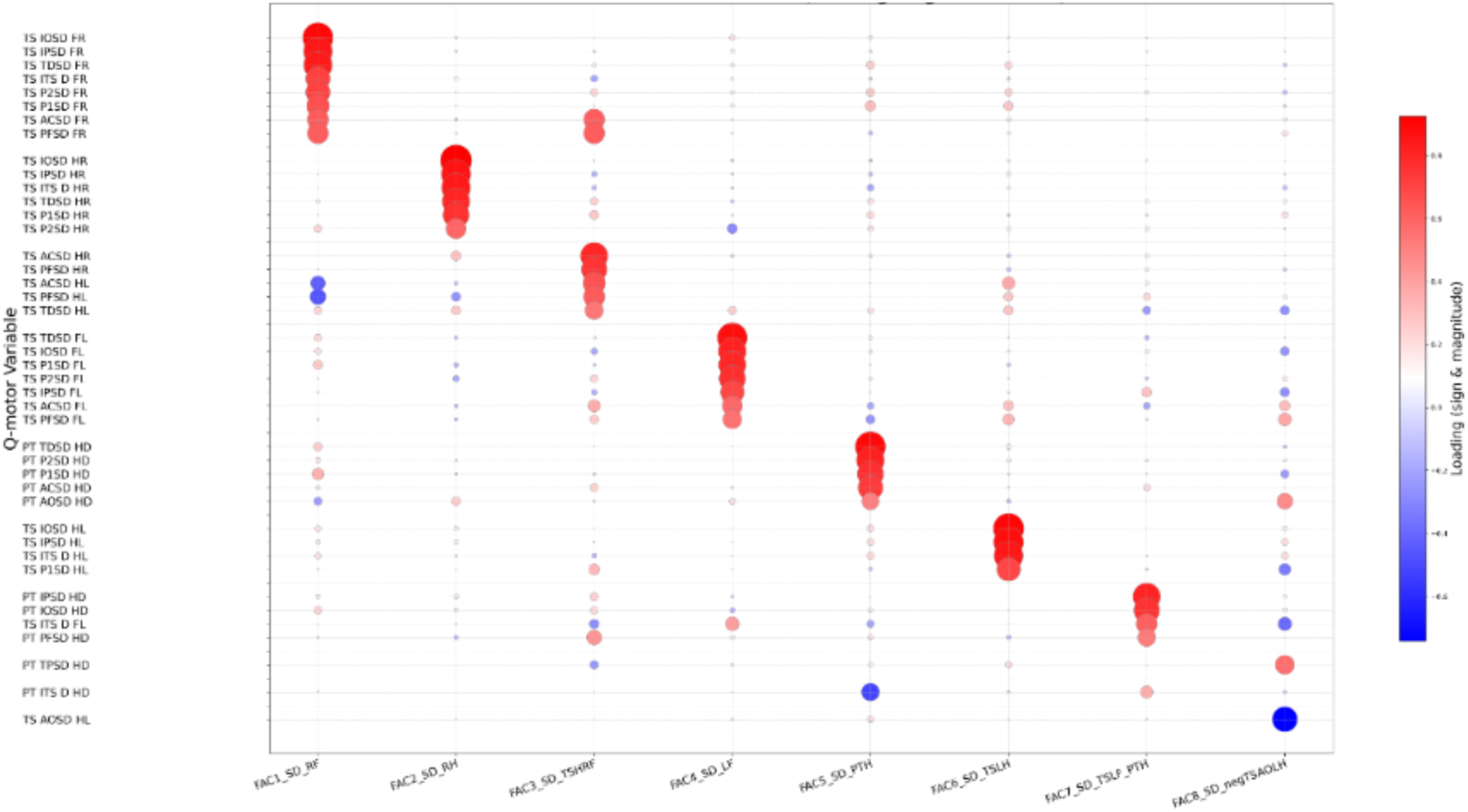
Bubble grid of component loadings for the Speeded Tapping and Pointing Variability PCA. Rows show “SD” variables included in the Speeded Tapping and Pointing Variability PCA and columns show retained principal components. Circle area is proportional to the absolute loading magnitude, and colour indicates loading sign and magnitude (red = positive; blue = negative), highlighting variables that contribute most strongly to each component.

**Figure S6.**
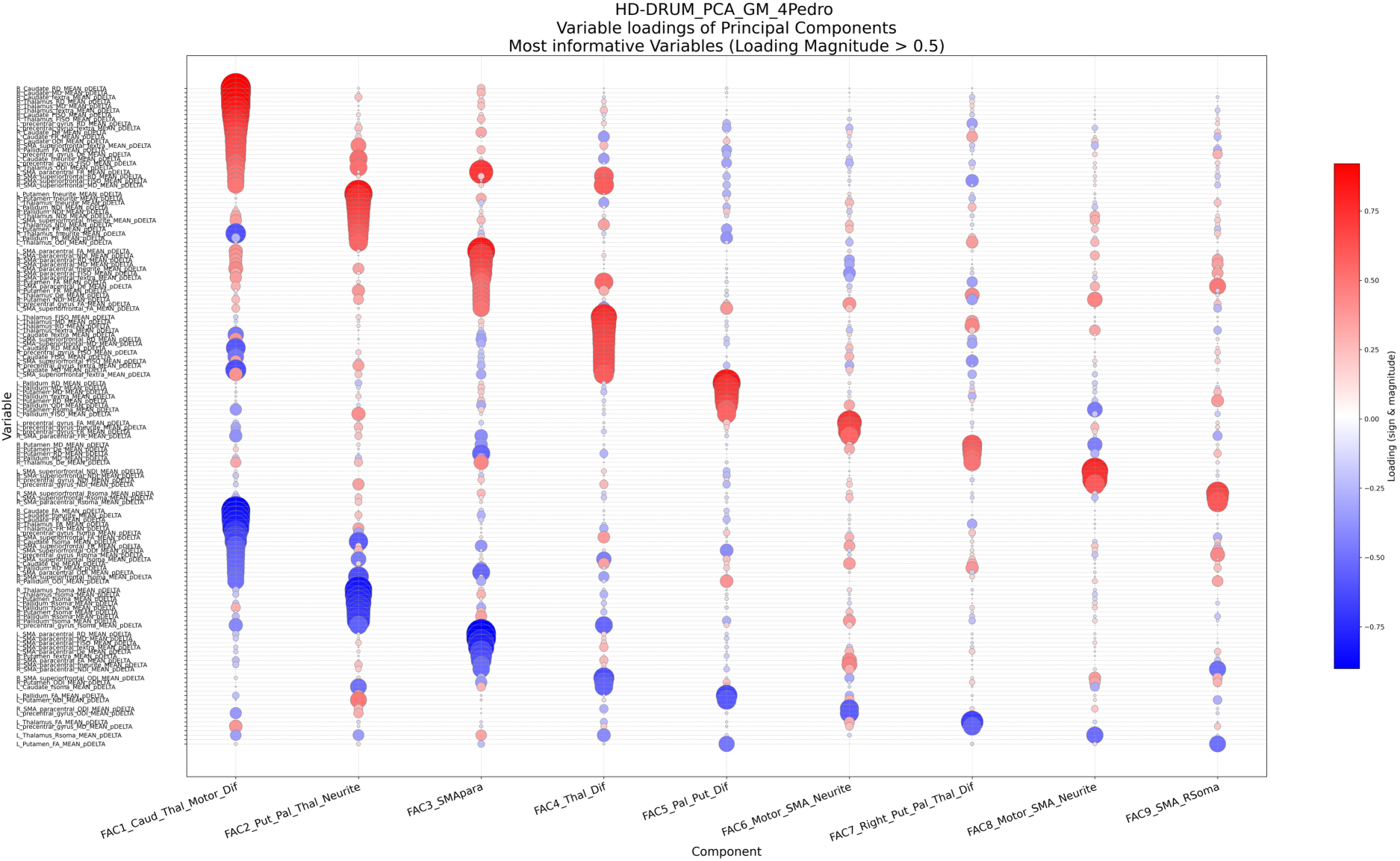
Bubble grid of component loadings for the grey matter microstructural PCA. Rows show the grey matter microstructural variables included in the PCA and columns show retained principal components. Circle area is proportional to the absolute loading magnitude, and colour indicates loading sign and magnitude (red = positive; blue = negative), highlighting variables that contribute most strongly to each component.

**Figure S7.**
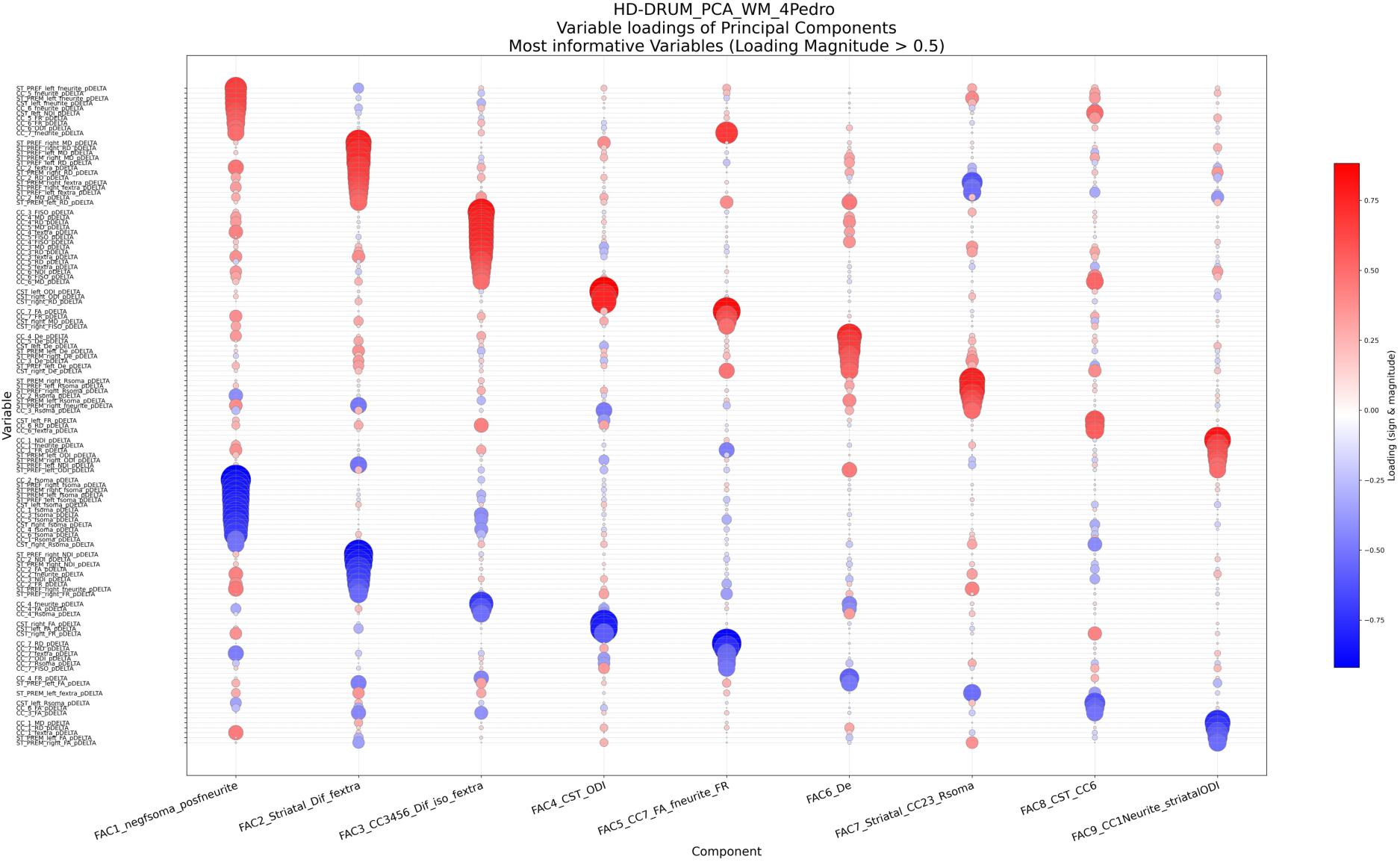
Bubble grid of component loadings for the white matter microstructural PCA. Rows show the white matter microstructural variables included in the PCA and columns show retained principal components. Circle area is proportional to the absolute loading magnitude, and colour indicates loading sign and magnitude (red = positive; blue = negative), highlighting variables that contribute most strongly to each component.

